# Self-Logical Consistency Assessment of Large Language Models for Patient Feedback Classification : Algorithm Development and Validation Study

**DOI:** 10.1101/2024.07.11.24310210

**Authors:** Zeno Loi, David Morquin, François-Xavier Derzko, Xavier Corbier, Sylvie Gauthier, Patrice Taourel, Emilie Prin-Lombardo, Grégoire Mercier, Kévin Yauy

**Affiliations:** Espace de Recherche et d’Intégration des Outils numériques en Santé, CHU Montpellier, Montpellier, France; Département des Maladies Infectieuses et Tropicales, CHU Montpellier, Univ Montpellier, Montpellier, France; Pôle soins, qualité, parcours et usagers, CHU Montpellier, Montpellier, France; Institut de Science des Données de Montpellier, Univ Montpellier, INSERM, Montpellier, France; Service d’Imagerie Médicale Lapeyronie, CHU de Montpellier, Montpellier, France; Institut Desbrest d’Épidémiologie et de Santé Publique, Univ Montpellier, INSERM, Montpellier, France; Unité de Recherche Médico-Économique, CHU Montpellier, Montpellier, France; Service de Génétique Médicale, LIRMM, CHU Montpellier, Univ Montpellier, Montpellier, France

**Keywords:** Artificial Intelligence, Natural Language Processing, Hallucinations, Quality of Health Care, Patient Satisfaction, Quality Improvement, Patient Rights, Classification, Reproducibility of Results, Health Services Research, Benchmarking, Models, Statistical, Feedback, Logic, Surveys and Questionnaires, Hospital Administration, Hospitals, University

## Abstract

**Background:** Patient satisfaction feedback is crucial for hospital service quality, but manual reviews are not possible due to their time-consumption, and traditional natural language processing methods remain inadequate. Large Language Models (LLMs) show promise but are prone to logical hallucinations—fabricated or illogical outputs that limit their reliability (inconsistent performance across repeated uses) and validity (explainability into clinical contexts) in healthcare.

**Objective:** This study aimed to evaluate the Self-Logical Consistency Assessment (SLCA), an original method designed to enhance LLM feedback classification reliability by enforcing a logically-structured chain of thought.

**Methods:** SLCA uses two validation steps: self-consistency (identifying the most coherent response) and logical consistency (ensuring alignment with the original statement and expert classifications). We evaluated SLCA using GPT-4 and Llama-3.1 405B on 12,600 classifications from 100 patient feedback samples to assess logical hallucinations, and tested its performance on a 49,140-classification benchmark derived from 1,170 feedbacks.

**Results:** SLCA reduced logical hallucinations among detected categories from 15.80% (168/1063) to 0.51% (4/786) with GPT-4 and from 7.17% (51/711) to 1.67% (10/599) with Llama-3.1, with residual errors confined to the emergency feedback category. On the benchmark, SLCA achieved precision-recall scores of 0.86-0.78 for GPT-4 and 0.84-0.58 for Llama-3.1. These results demonstrate SLCA’s ability to achieve human-level performance across LLMs.

**Conclusions:** SLCA offers a zero-shot, scalable, explainable solution for improving LLM classification reliability in healthcare. Its capacity to enhance performance without fine-tuning positions it as a valuable tool for analyzing patient feedback and supporting hospital service quality improvement.

*What is already known on this topic:* Free-text patient feedback labeling is crucial for healthcare system improvement. Classification by hand is impractical due to its time-consumption. Large language models (LLMs) can out-perform traditional NLP for this task, but their clinical use is limited by inconsistent predictions and “logical hallucinations” that undermine explainability and trust.

*What this study adds:* The Self-Logical Consistency Assessment (SLCA) framework, which couples self-consistency with a novel logical-consistency check, almost eradicates hallucinations (from 15.8 % to 0.5 % with GPT-4) while reaching human-level precision (86%) and better exhaustivity (recall +14%).

*How this study might affect research, practice or policy:* SLCA offers a scalable, explainable and data-sovereign pathway for hospitals and regulators to adopt LLMs in routine patient-experience monitoring, and it provides a transferable template for evaluating AI safety in other clinical-text tasks.

**Graphical Abstract:** Graphical abstract

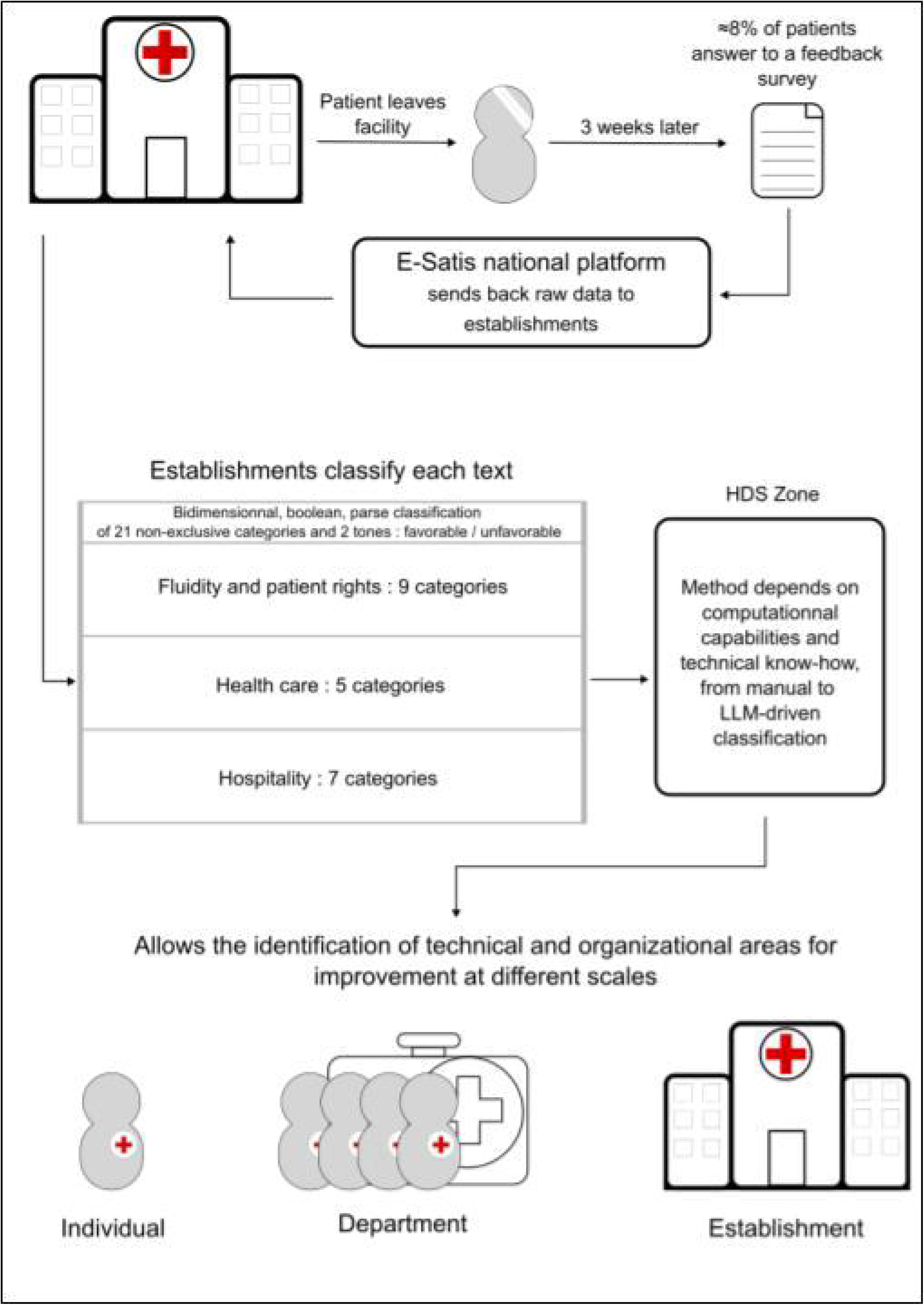

## INTRODUCTION

Patient satisfaction feedback is a crucial metric for determining areas of improvement in hospital services, directly impacting quality of care [1][2]. In French hospitals, patient feedback is collected systematically through multiple channels to capture diverse patient experiences. Feedback is primarily gathered via the national e-Satis survey, administrated by the French National Health Authority (HAS), which is emailed to patients three weeks after discharge if consent is provided. Additionally, institutional satisfaction questionnaires are completed by patients before discharge, and spontaneous patient communications—either complaints or expressions of satisfaction—are received through the hospital’s Patient Relations Department (Figure 1).

**Figure 1 :**
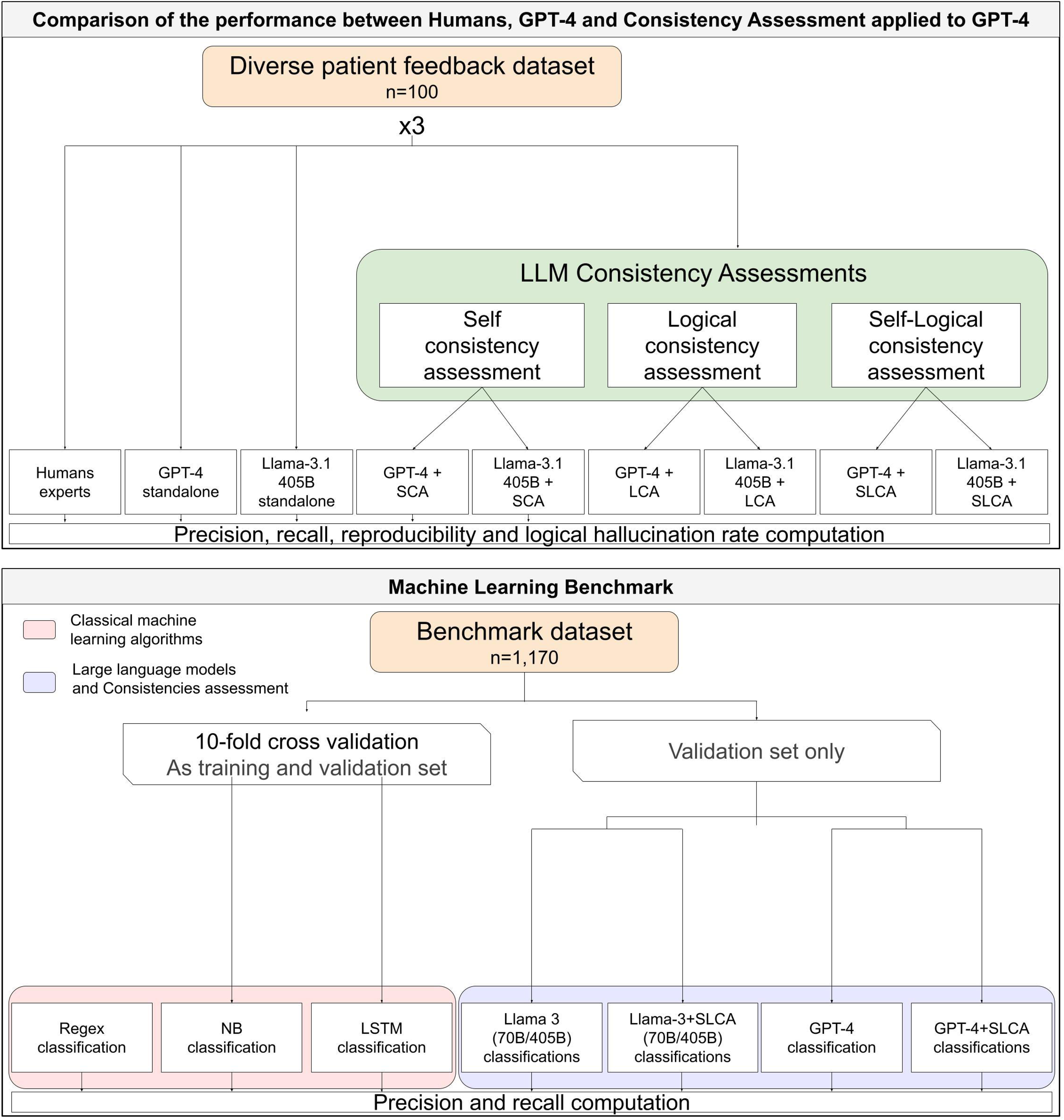
Computer and clinical processing of patient feedback

Patient feedback offers clinical and organizational insights in three domains—Fluidity & Patient Rights, Health Care, and Hospitality. Sub-topics span for example emergency-service responsiveness, staff empathy and availability, patient rights, and comfort factors such as room temperature. Precise topic labeling lets staff target improvement priorities, but manual classification is too time-consuming for large-scale deployment.

Montpellier Regional Hospital analyzes feedback with a regex-based keyword dictionary in Sphinx to produce the quarterly “Patient Experience Barometer.” Even with periodic updates, the method’s reliance on fixed terms overlooks nuanced, context-rich narratives and complex clinical sentiments. Like other legacy NLP approaches, it needs replacement by a model that can grasp semantically rich, clinically relevant content. In 2022, an unsupervised classification of 2.5 million feedbacks provided a comprehensive overview of patient concerns and defined 20 categories [3,4]. However, models like Naive Bayes, BERT and convolutional networks struggle to accurately classify nuanced feedback because they cannot effectively handle complex language contexts [5–7][6].

Large Language Models (LLMs) like GPT-4 and Llama-3.1 405B offer a promising alternative due to their superior ability to understand natural language and detect subtle nuances in patient feedback [6,8],[9]. Moreover, they might offer the needed explainability of their classifications, which is mandatory in high stakes language processing[10]. They have been recently applied in exploratory studies for quality measurement purposes [11] and specific language handling tasks in healthcare[12], including text classification[13]. However, since these models have been shown to exhibit inaccuracies in the medical field [14], there is a pressing need to improve the reliability of text classification. Recent work shows that evaluating the self-consistency of LLM predictions — that is, ensuring the model provides the same classification over multiple independent attempts — greatly enhances performance in classification tasks [15]. However, these models often produce hallucinations, which are factual mistakes or logical flaws in generated text [16]. Although minor factual hallucinations may be tolerable in certain hospital contexts, logical hallucinations—also known as “extrinsic faithfulness hallucinations”—introduce illogical reasoning that undermines explainability and is therefore incompatible with sensitive healthcare applications. While these logical hallucinations directly impact reliability in a critical way, they also clearly jeopardize the explainability of resulting classifications[17]. Ethically, healthcare choices must be traceable from data collection to decision, grounded in explainable, verifiable evidence. When a patient-experience barometer presents nonsensical categories, clinicians lose confidence, breaking the feedback loop with patients. Because AI lacks the interpersonal trust granted to humans, it must offer explicit, transparent explanations for its outputs[18]. Since Chain of Thought (CoT) generation methods improve the explainability of these models [19], validating CoT logical structures might be a potential solution to the issue of logical hallucinations.

To address this challenge, we developed Self-Logical Consistency Assessment (SLCA), a method designed to improve the reliability of LLM-generated predictions. The SLCA combines Self-Consistency Assessment (SCA) [15] — the evaluation of the LLM ability to provide the same predictions in similar conditions — together with an original method called Logical Consistency Assessment (LCA), which appraises the capability of an LLM to produce a logically structured CoT. The aim of this study is to evaluate whether SLCA can achieve human-level performance and provide a practical option in the automated classification of patient feedback.

## METHODS

### Data Collection and Samples

From 2022 – 2024, 1,270 adult-patient feedbacks were retrieved from the national E-Satis system for Montpellier University Hospital. We excluded records from patients who refused data use, messages too long for analysis, compensation claims, and feedback with extreme content (Supplementary Figure 1). All retained entries were pseudonymized, and none were removed during the study.

Two datasets were built:

- **Human-Curated sample (n = 100):** Selected for feedback diversity to compare human-versus GPT-4-based consistency assessments (Supplementary Table 2). Three raters of each type classified this set to measure reproducibility.
- **Benchmark sample (n = 1,170):** Randomly drawn from 2023 to benchmark models (Supplementary Table 3). Power analysis sought to detect a 2 % precision or recall difference across 16 tests with α = .01 (Bonferroni) and 1 − β = .80. This required 45,600 classifications—1,086 feedbacks—so we included 1,170 to maintain power (Supplementary Table 5).

### Patients and public involvement

Patients contributed only at the data-collection stage, as the feedback were voluntarily submitted to the national e-Satis survey after discharge during 2022-2024. Their aggregated preferences shaped the research question, because the 21-category framework we tested was adapted from the Haute Autorité de Santé topic-model analysis of 2.5 million French patient narratives[3]. No patients or public representatives took part in protocol design, category definition, results interpretation or manuscript review. The study imposed no additional burden on respondents and no burden assessment was undertaken, as comments were supplied within routine feedback infrastructure.

### Categorisation Approach

The classification task involved categorizing feedback into 21 non-exclusive categories and two non-exclusive tones—favorable and unfavorable (Figure 1 and Supplementary Note 1). The categories were adapted from the Haute Autorité de Santé (French National Health Authority) guidelines [3,4], with the addition of “Patient’s Rights” to meet local healthcare quality improvement goals. The favorable tone describes positive aspects appreciated by patients, while the unfavorable tone indicates criticisms or disliked aspects.

### Evaluation metrics

Comparisons focused on:

- Precision: The proportion of relevant items among those selected, essential for medical-grade classification.
- Recall: The proportion of relevant items that were selected.
- Reproducibility: Assessed via agreement between different agents.
- Hallucination Rate: Following prior work [16], logical hallucinations occur when the LLM generates a false positive from information not inferable from the original text or includes unjustified logical steps, disabling the explainability of the prediction. Logical hallucinations were assessed by cross-referencing the LLM’s output with patient feedback, verifying that all information in the CoT leading to a category identification was explicitly stated or inferable. If the CoT included additional information or logical inconsistencies, it was flagged as containing a logical hallucination. The hallucination rate is defined here as the percentage of logical hallucinations among argued category identifications. The benchmark focused on precision and recall for overall performance; subgroup analyses used the F1 score.

### Logical Consistency Methodology

We developed SLCA by combining two complementary methods: Self-Consistency Assessment (SCA) and Logical Consistency Assessment (LCA). SCA focuses on reproducibility: the language model is prompted two times for the same feedback, and only categories that appear consistently across runs are retained. This helps reduce sporadic or inconsistent classifications. Despite its efficiency to enhance precision, SCA does not address the explainability flaw LLMs tend to present when they produce logical hallucinations. Evaluating logical human argumentation is fundamental in the philosophy of logic [20]. While prior studies structured LLMs’ Chains of Thought (CoT) [21],[22], none directly applied philosophical methodologies to assess logical validity in LLM-generated CoTs. Our work integrates these methods to enhance the logical consistency of LLM predictions.

LCA evaluates whether the model’s reasoning is structurally valid. Specifically, each classification is broken down into:

- **Premise:** A quote from patient feedback.
- **Implication:** The logical link connecting the premise to the conclusion.
- **Conclusion:** A category identification.

An argument is valid only when its premise logically entails the conclusion. For each of the 21 feedback categories, we assembled a domain-specific implication set agreed upon by three human experts and the Principal Investigator. The LLM must justify every label with one approved premise-implication-conclusion chain; we then verify that the LLM is able to produce a structure-valid CoT, based on the predefined set of implications. Example: “The nurse was incredibly kind and attentive” legitimately supports “Humanity and Availability of Professionals.” If the model selects this category with an invalid implication, such as “Quality and Speed of Response from Calls to Regulatory Services,” LCA flags a logical hallucination. Combining this logic filter with SCA reproducibility forms SLCA, which boosts consistency, explainability, and overall reliability

### Comparing LLMs and human classification

Figure 2 represents the study design on the two produced analyses, the first being explainability oriented and the second being precision-recall focused.

**Figure 2 :**
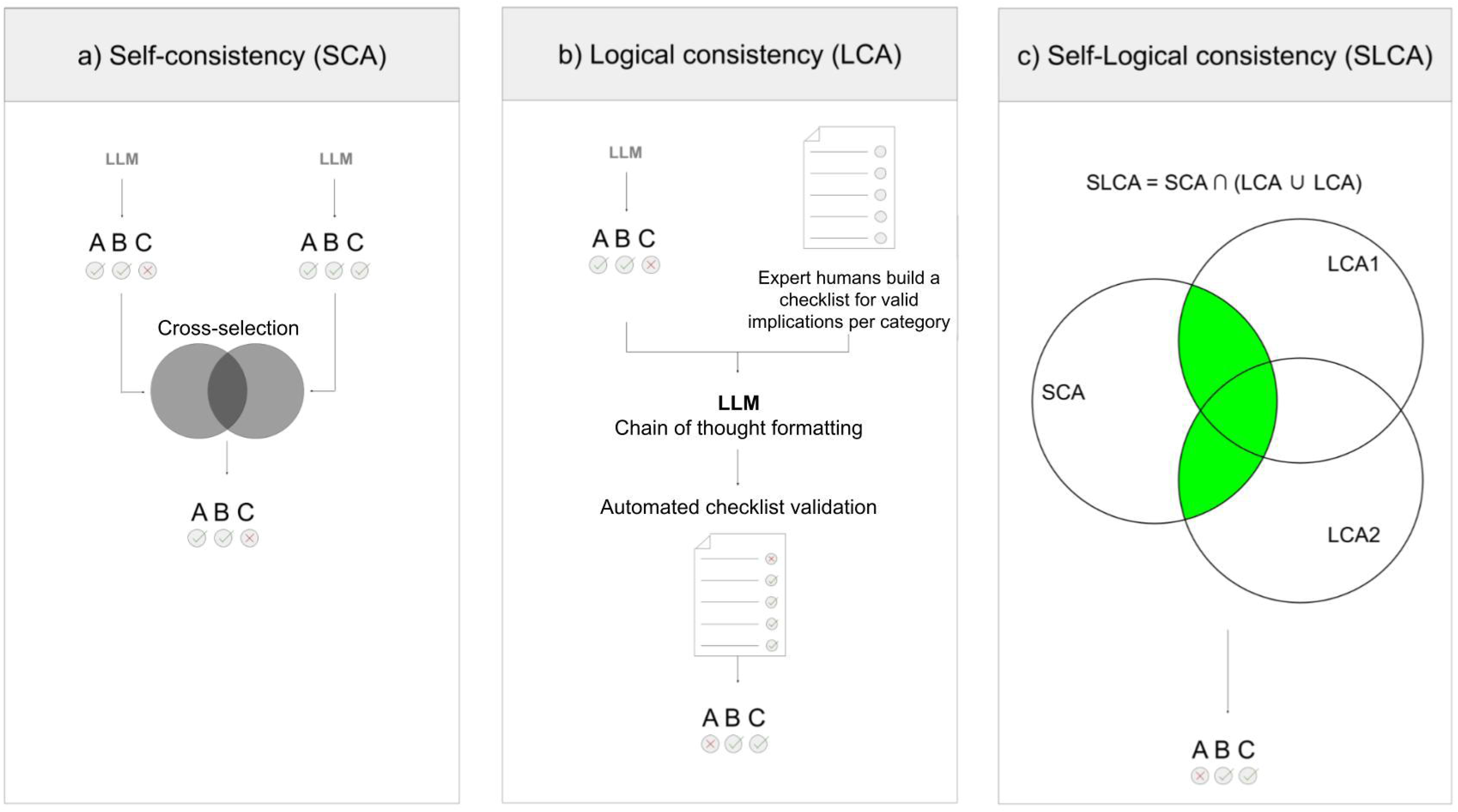
Consistency Assessment Evaluation Study Design

This section analyses are led on the 100 feedbacks human-curated sample (Figure 2).

- **Human Expert Evaluation:** Three quality-of-care experts independently classified each feedback. No communication occurred between them. A fourth expert blindly reviewed each classification to establish the gold standard (Supplementary Table 4).
- **LLMs Classification Methods:**

Both GPT-4 turbo (version March 05, 2024) and Llama-3.1 405B were evaluated independently as raw predictive models and in their consistency assessed versions as following :

○ **Standalone LLM :** Three runs of LLMs classified each feedback independently. The prompt included output structure, classification conditions, category list, and instructions to generate a structure-free CoT (Supplementary Note 2). No example was given.
○ **Self-Consistency Assessment (SCA):** Three additional LLMs runs classified each feedback independently. Classifications were grouped in three pairs. For each GPT-4+SCA agent, only categories identified by both runs were retained, aiming to select consistently identified categories.
○ **Logical Consistency Assessment (LCA):** To assess logical consistency, an initial prediction was produced without CoT structure restrictions. A second prompt instructed the LLM to structure its previous output into a logical argument (Supplementary Note 2), using implications from the predefined list (Supplementary Note 1). For each LLM+LCA agent, only classifications with a valid implication were retained.
○ **Combined Self-Logical Consistency Assessment (SLCA):** Both self and logical methods were applied, selecting identifications fulfilling two conditions: both runs identified the category, and at least one produced a valid CoT. This process evaluates the LLM’s logical consistency and self-consistency, enhancing precision and exhaustivity.

Figure 3 presents the LLM call patterns and post-processing in the three consistency assessment methods. SLCA does not require additional LLM inferences after LCA and SCA as it is deduced by logical combination.

**Figure 3 :**
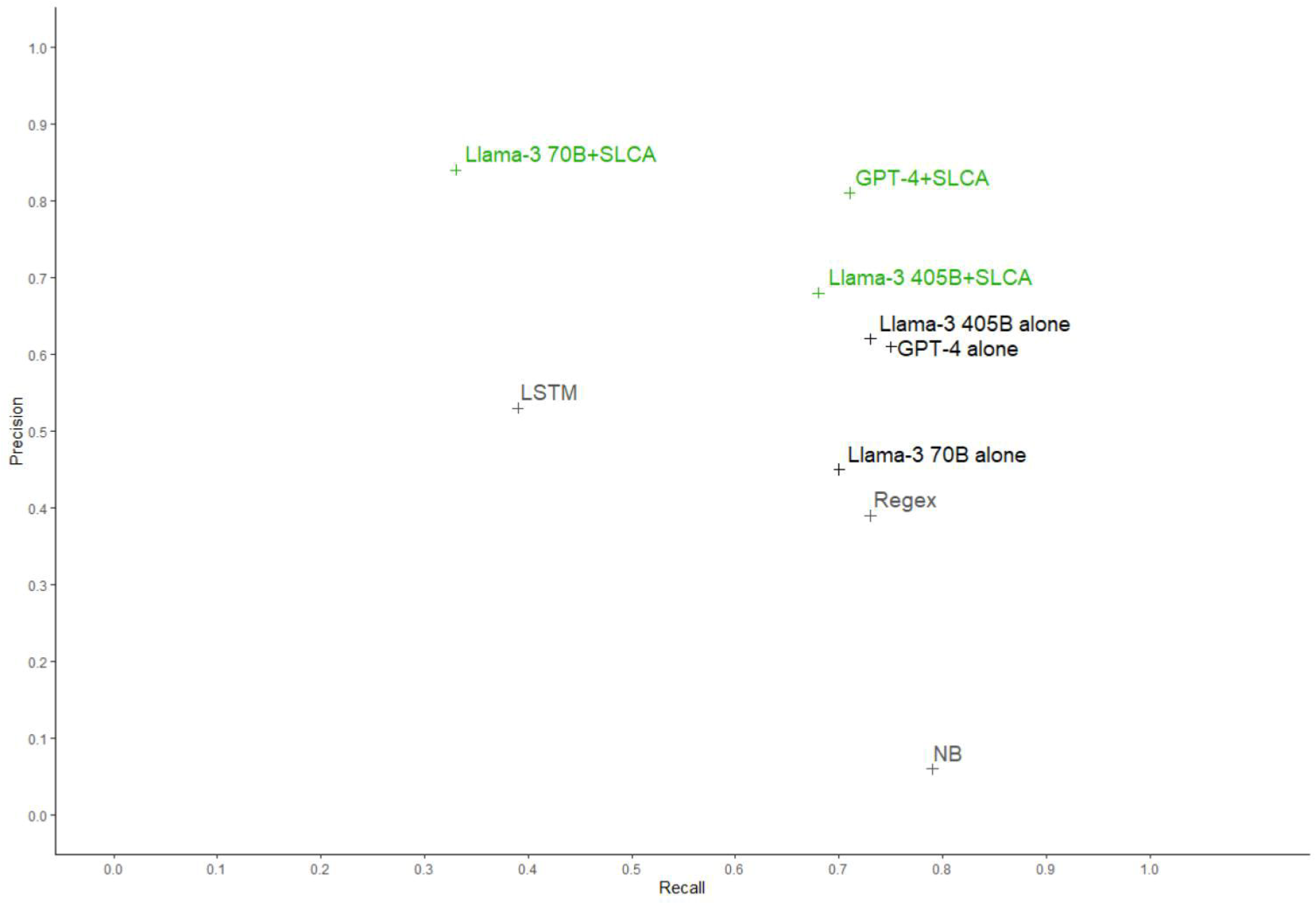
Consistency Assessment Layouts

### Machine Learning Benchmark

The benchmark compared 11 models over 49,140 classifications among 1,170 feedbacks (Figure 2) :

- **Traditional Models:** Naive Bayes (NB) and a 1.5 million parameter Long Short-Term Memory (LSTM), trained and evaluated using ten-fold cross-validation.
- **Regex-Based Model:** Used in its production version at our facility.
- **Llama-3.1 70B and Variations:** Standalone and consistency-assessed variations (Llama-3.1 70B+SCA, Llama-3.1 70B+LCA, Llama-3.1 70B+SLCA).
- **Llama-3.1 405B and Variations:** Standalone and consistency-assessed variations (Llama-3.1 405B+SCA, Llama-3.1 405B+LCA, Llama-3.1 405B+SLCA).
- **GPT-4 Turbo and Variations:** Standalone and variations (GPT-4+SCA, GPT-4+LCA, GPT-4+SLCA).

NB was performed after dimensional reduction using non-negative matrix factorization. Due to time constraints, the gold standard for this experiment was created by a single quality-of-care expert.

### Statistical analysis

All classification outputs were treated as categorical data (category present vs. absent). For the human-curated sample (100 free-text feedbacks), each feedback was independently analyzed three times by each agent (human, LLM variations), resulting in 12,600 binary classifications (21 categories plus 2 tones per feedback). Precision, recall, and hallucination rates were compared using McNemar tests (two-tailed α=.05) on pairwise differences between models. Krippendorff’s alpha was computed to assess reproducibility among agents of the same type. For the benchmark dataset (1,170 feedbacks, yielding 49,140 binary classifications), overall performance metrics (precision, recall) were compared by McNemar tests (two-tailed α=.05) on pairwise differences between models. Confidence intervals (95% CIs) for precision and recall were derived using the Wilson method. All analyses were performed using the R (version 4.0) statistical environment.

### Ethical Considerations

This study complies with French data protection laws. Patients were informed about data usage and could withdraw access at any time. Ethical approval was granted by the Ethical and Scientific Committee of the Montpellier University Hospital Centre (Registration Number: A015/2024-05-050/001).

## RESULTS

### Comparing LLMs and human classification

We compared the performance of LLMs (GPT-4 and Llama-3.1 405B) to human experts in classifying 12,600 categories across 100 patient feedback samples. We evaluated precision, recall, time efficiency, and hallucination rates. The feedback was diverse, with all categories represented. The three most prevalent categories were “Humanity and availability of professionals”, “Medical and paramedical care” and “Facilities and rooms”, consisting in 50% of all categories to identify.

Humans demonstrated high precision (0.87) but low recall (0.64), reflecting their cautious approach and tendency to underclassify. Classification was also time-intensive, requiring approximately 3 hours to process 100 feedback samples, underscoring the inefficiency of manual reviews for large-scale analyses.

LLMs were slightly less precise than humans, with GPT-4 achieving a precision of 0.71 and Llama-3.1 405B achieving 0.76. However, LLMs were significantly more exhaustive, with GPT-4 achieving a recall of 0.88 compared to 0.63 for Llama-3.1 405B (McNemar compared to humans respectively : χ²=286, df=1, *P*<.001 for GPT-4 and χ²=12, df=1, *P*<.001 for Llama-3.1 405B). These findings highlight the potential of LLMs for identifying more relevant categories while sacrificing some precision.

A major limitation of LLMs was their susceptibility to logical hallucinations. GPT-4 exhibited a hallucination rate of 15.67% (168/1063), while Llama-3.1 405B showed a lower rate of 7.17% (51/711). These logical hallucinations represent fabricated or misaligned category identifications, undermining the reliability of raw LLM outputs without additional safeguards.

We progressively evaluated the effects of Self-Consistency Assessment (SCA), Logical Consistency Assessment (LCA), and Self-Logical Consistency Assessment (SLCA) on GPT-4 and Llama-3.1 405B. Evaluation was focused on their impact on precision, recall, hallucination rate reduction, and reproducibility.

SCA significantly improved precision for both LLMs, increasing GPT-4’s precision by 12% (McNemar χ²=209, df=1, *P*<.001) and Llama-3.1 405B’s by 7% (McNemar χ²=92, df=1, *P*<.001). However, hallucination rates remained significant, with GPT-4+SCA exhibiting a 5.87% (50/852) hallucination rate and Llama-3.1 405B+SCA showing 5.19% (32/617). GPT-4+SCA achieved a precision-recall score of 0.83-0.83, while Llama-3.1 405B+SCA showed slightly lower recall with a score of 0.83-0.59 (McNemar χ²=129, df=1, *P*<.001). These results indicate that while SCA enhances precision, it does not adequately address hallucination rate issues.

LCA further improved performance by addressing logical hallucinations. GPT-4+LCA reduced hallucination rates to 1.99% (18/903), achieving a precision-recall score of 0.76-0.80, though it remained less precise and more exhaustive than human evaluations (McNemar χ²=131, df=1, *P*<.001). Llama-3.1 405B+LCA achieved a precision of 0.77 and a recall of 0.63, with hallucination rates reduced to 2.27% (16/704). Notably, all logical hallucinations for both models were confined to the “Medical and Paramedical Care” category, specifically related to “Quality and Speed of Response from Calls to Regulatory Services and Emergency Services (EMS, emergency department).”

SLCA combined SCA and LCA for maximal performance gains. GPT-4+SLCA achieved human-equivalent precision of 0.86 and a greater recall of 0.75, (McNemar χ²=53, df=1, *P*<.001), with hallucination rates reduced to just 0.51% (4/786). Llama-3.1 405B+SLCA exhibited slightly lower performance with a precision-recall score of 0.84-0.58 and a hallucination rate of 1.67% (10/599) (McNemar χ²=87, df=1, *P*<.001). Importantly, logical hallucinations for both models occurred exclusively under the same identifiable circumstances noted with LCA.

SLCA improved inter-rater agreement, with Krippendorff’s alphas of 0.86 for GPT-4+SLCA and 0.87 for Llama-3.1 405B+SLCA, compared to 0.67 for human experts. This indicates that SLCA not only enhances classification performance but also ensures greater consistency.

### Machine Learning Benchmark

The external validation of 49,140 classifications among 1,170 feedbacks was quantitatively representative with each categories represented several times. The three most prevalent categories were “Humanity and availability of professionals”, “Speed of care and waiting time” and “Meals and snacks”, consisting in 48% of all categories to identify. This benchmark revealed a clear progression in performance, from traditional methods to state-of-the-art LLMs enhanced with SLCA, in terms of precision and recall (Figure 4).

**Figure 4 :**
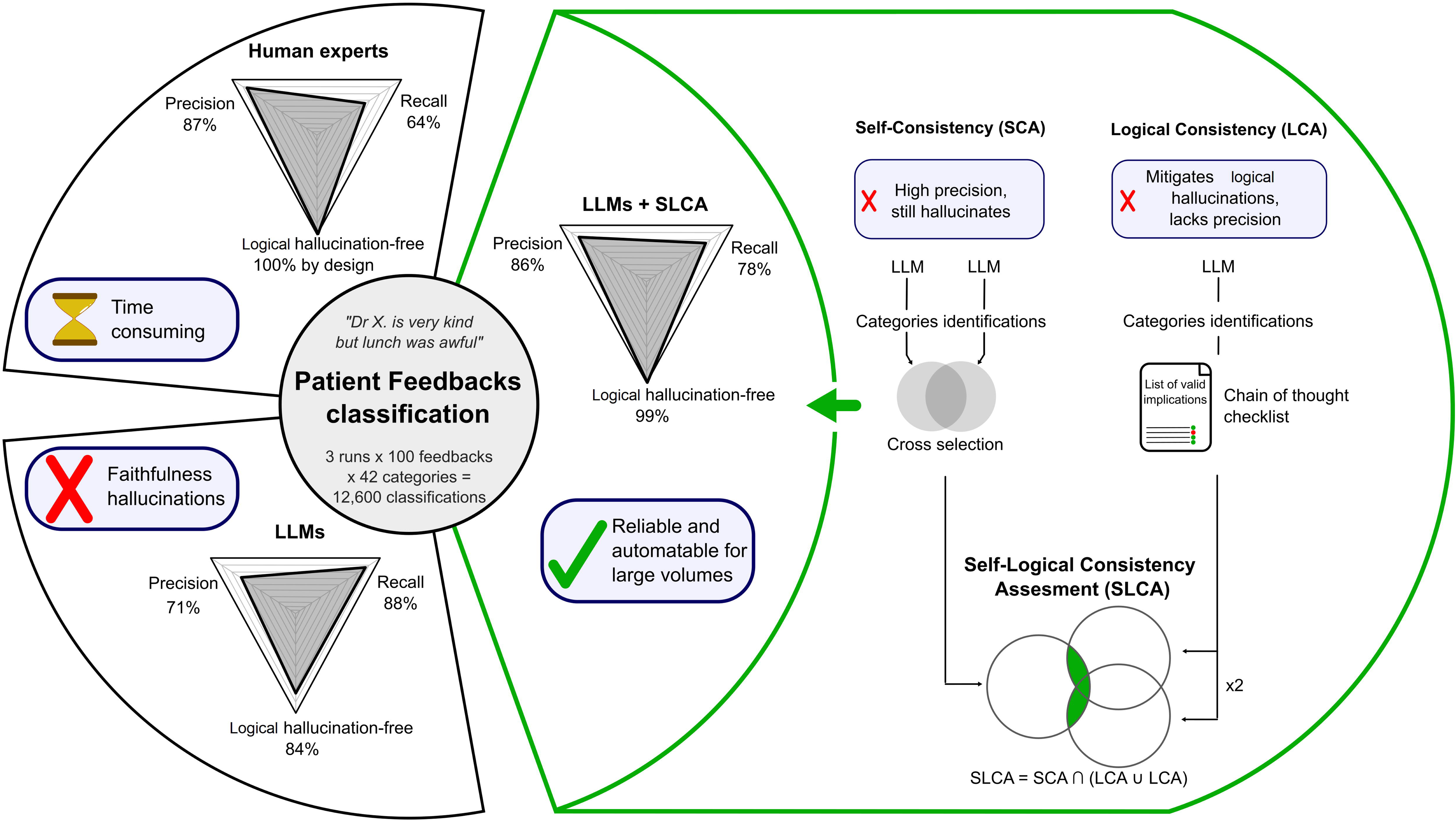
Benchmark of Machine Learning Models on Patient Feedback Classification

Historical methods demonstrated limited capabilities. Regex achieved a precision-recall score of 0.39-0.73, performing reasonably well in recall but struggling with precision. LSTM, while slightly more precise at 0.53, had a recall of only 0.39, reflecting its inability to capture the full complexity of patient feedback. Naive Bayes performed worst, with a precision of 0.06 despite a recall of 0.79, indicating a significant overclassification issue.

Standalone Large Language Models (LLMs) showed considerable improvement over historical models. GPT-4 exhibited a recall of 0.75 but suffered from poor precision at 0.61, highlighting a tendency to overclassify. Similarly, Llama-3.1 405B performed comparably with precision-recall scores of 0.62-0.73. Llama-3.1 70B, the smallest of the LLMs tested, underperformed, achieving a low precision of 0.45 despite an acceptable recall of 0.70.

SLCA markedly enhanced LLM performance. GPT-4+SLCA achieved the highest precision (0.88) and recall (0.71). Llama-3.1 405B+SLCA followed with balanced precision-recall scores of 0.68-0.68. While Llama-3.1 70B+SLCA exhibited high precision (0.82), its recall remained limited at 0.30, highlighting challenges in comprehensiveness for smaller models. The progression from traditional models to standalone LLMs and ultimately to SLCA-enhanced LLMs demonstrates a transformative leap in classification reliability.

All differences between models predictions were statistically significant (McNemar χ²∈[3.85;20536] across 36 pairwise comparisons, df=1 *P*<.05).

The benchmark subgroup analysis reveals notable variations in model performance across patient feedback categories.

Certain categories consistently posed challenges for all evaluated models. For example, “Humanity and Availability of Professionals — favorable” and “Medical and Paramedical Care — favorable” exhibited relatively lower F1 scores, ranging from 0.76 to 0.94. These categories likely involve nuanced or subjective interpretations, making them difficult for automated systems to classify accurately.

In contrast, categories such as “Room Temperature — favorable” and “Patient Rights — favorable” achieved near-perfect performance, with F1 scores between 0.99 and 1.00. These categories are characterized by more straightforward, objective feedback, making them easier for models to process effectively. The stark difference between these and more challenging categories highlights the variability in model performance.

The analysis identified four outlier categories (out of 42) that were either particularly challenging or notably easier to classify. Importantly, this trend was consistent across all models, including both standalone and SLCA-enhanced LLMs. Notably, logical hallucinations identified in the earlier LCA evaluation were confined to the challenging category “Medical and Paramedical Care.” These findings illustrate the evolving performance landscape across patient feedback categories, from highly reliable classifications in straightforward categories to persistent challenges in nuanced ones.

## DISCUSSION

### Key results

Human experts labeled feedback accurately but missed many cases and required substantial time. With SLCA, the LLM reached human-level precision, gained recall, kept hallucinations rare, and showed stronger reproducibility in external tests. This explainable workflow makes large-scale, human-grade, feedback classification feasible for systems requiring explainable inference like those in France and the United Kingdom.[10,23].

### Strengths

The SLCA framework is built on a series of nested models : the LLM alone, SCA, and LCA. Each step increases precision or explainability at the expense of recall, offering flexibility to adjust accuracy and coverage based on clinical needs. Our method sets a new state of the art for patient feedback classification. While the best previously reported model, a BERT variant, achieved 70% accuracy [6], GPT-4 and Llama-3.1 405B with SLCA reached respectively 98% and 96%, thereby illustrating a substantial improvement over existing approaches. The Montpellier University Hospital Centre already implements this method to leverage patient feedback.

Additionally, SLCA nearly eliminates logical hallucinations. In 100 feedback entries, only 33 GPT-4 and 35 Llama-3.1 405B classifications (of 12 600 total) involved the “Quality and Speed of Response from Calls to Regulatory Services and Emergency Services (EMS, emergency department)” implication, yielding the 4 GPT-4 and 10 Llama-3.1 405B hallucinations observed. With so few cases, human review is straightforward. Although performance on larger datasets remains to be tested, the results are clinically promising, and Llama-3.1 405B offers a sovereign alternative where proprietary GPT-4 use is constrained.

Also, SLCA-enhanced LLMs reach higher recall than human reviewers without sacrificing precision. They supply written justifications that sharpen category definitions and enable finer, incremental process improvements. Humans, though considered hallucination-free, cannot feasibly record a rationale for every judgment.

Each SLCA prediction needs four LLM inferences, increasing compute cost, yet it adds crucial agility: models and label schemes can be updated fast without fine-tuning or re-annotation, easing human bottlenecks. Large-scale deployment may still call for extra speed optimizations, but the overhead is outweighed by its adaptability, explainability, performance gains, and reduced manual upkeep.

Finally, gold standards shift, so inter-expert agreement is weak and consensus elusive— exactly the challenge in real-world NLP evaluation. By matching human precision and recall, our method supplies a stable, more reproducible reference and paves the way for AI-generated gold standards when manual ones are impractical.

### Limitations

Smaller models (e.g., Llama-3.1 70B) underperformed, showing that SLCA’s effectiveness depends on base-model capacity and may hit scalability limits in resource-constrained hospitals. We evaluated SLCA alone, but pairing it with other consistency checks could further lift performance. A major limitation was our inability to inspect model internals—an approach recent work uses to cut hallucinations—leaving room for future gains. [24],[25].

Clear, precise categories matter: broad labels such as “medical and paramedical care” caused inconsistent results, urging sharper definitions. Future upgrades include (1) expanding the dataset to validate SLCA externally and reveal factual hallucinations, (2) cross-validating logical-hallucination flags with multiple investigators to curb subjectivity, and (3) testing language effects, since the corpus is French and LLMs often excel in English.

### Conclusions

Overall, SLCA presents practical benefits in healthcare, where patient feedback is crucial for quality improvement ^3,12^. Its ability to achieve high accuracy with greater control over logical hallucinations and control over computational cost while saving the possibility of complete sovereignty of the patients data makes it a valuable tool for large-scale feedback processing. This advancement provides healthcare institutions with more accurate, clinically meaningful insights from patient narratives, facilitating targeted quality improvements and enhancing the reliability of automated clinical informatics analyses. As this method is not specifically designed for patient feedback classification, assessing consistency might be all you need to classify complex text data with explainability and precision. For all healthcare structures, from establishments to national health authorities, transitioning from classic NLP solutions — which requires regular manual updates and offers limited semantic nuance— to SLCA-enhanced LLM classifications would significantly improve the accuracy and granularity of clinical informatics extracted from patient feedback. Furthermore, this study serves as a preliminary exploration of generative AI’s reliability in structuring more critical health data, where maintaining logical consistency is all the more essential. Such a transition would require an initial technological investment to integrate the SLCA results into the existing software interfaces, yet this investment could yield rapid returns due to decreased time spent on feedback processing and increased quality of insights provided to frontline healthcare providers.

**Table 1 :** Patient Feedback Classification Performances of Humans, LLM and Consistency Applied to LLM.

| Agents | Mean performances over three independent agents |  |  | Classification reproducibility (Krippendorff's alpha) |
| --- | --- | --- | --- | --- |
|  | Precision (%) | Recall (%) | Hallucination rate (%) |  |
| Human experts | 87 | 64 | 0 (by design) | 0.67 |
| GPT-4 standalone | 71 | 88 | 15.80 (168/1063) | 0.81 |
| GPT-4 + SCA | 83 | 83 | 5.87 (50/852) | 0.87 |
| GPT-4 + LCA | 76 | 80 | 1.99 (18/903) | 0.82 |
| GPT-4 + SLCA | 86 | 78 | 0.51 (4/786) | 0.86 |
| Llama-3.1 405 B standalone | 76 | 63 | 7.17 (51/711) | 0.87 |
| Llama-3.1 405 B + SCA | 83 | 59 | 5.19 (32/617) | 0.89 |
| Llama-3.1 405 B + LCA | 77 | 63 | 2.27 (16/704) | 0.86 |
| Llama-3.1 405 B + SLCA | 83 | 59 | 1.67 (10/599) | 0.87 |

## Supporting information

TRIPOD-LLM Checklist

## Data Availability

Datasets are available from the corresponding authors on reasonable request. Pseudonymized patient feedback may contain personal health information; access is granted with traceability according to French National Commission on Informatics and Liberty and Montpellier University Hospital Centre policies.

https://github.com/ERIOS-project/SLCA_LLM4Quality

## Abbreviations

AI: Artificial Intelligence
BERT: Bidirectional Encoder Representations from Transformers
CHU: University Hospital Center (from the French “Centre Hospitalier Universitaire”)
CoT: Chain of Thought
EMS: Emergency Medical Services
ERIOS: Research and Integration Unit for Digital Tools in Healthcare (from the French “Espace de Recherche et d’Intégration des Outils numériques en Santé”)
F1: F1 Score, the harmonic mean of precision and recall
GPT-4: Generative Pre-trained Transformer 4
HAS: French National Health Authority (from the French “Haute Autorité de Santé”)
INSERM: French National Institute of Health and Medical Research (from the French “Institut National de la Santé et de la Recherche Médicale”)
ISDM: Montpellier Data Science Institute (from the French “Institut de Science des Données de Montpellier”)
LCA: Logical Consistency Assessment
LIRMM: Montpellier Laboratory of Informatics, Robotics, and Microelectronics (from the French “Laboratoire d’Informatique, de Robotique et de Microélectronique de Montpellier”)
LLM (or LLMs): Large Language Model(s)
LSTM: Long Short-Term Memory
NB: Naive Bayes
NLP: Natural Language Processing
SCA: Self-Consistency Assessment
SLCA: Self-Logical Consistency Assessment
95%□CI: 95% Confidence Interval

## Code and Data Availability

The code is available on GitHub as presented in references [26].

## Acknowledgments

The study was conceived, funded, and executed entirely by CHU de Montpellier. We acknowledge all participants and professionals involved in the patient feedback processing. Special thanks to the ERIOS team: Cécile Yriarte, Louise Robert, Laurine Moniez, Quentin Luzurier, Marin Portalez, Letizia Pala, Loïc Fontaine, Mylene Fernandes, Rita Pires, Yrina Gilhodes, Anne Laurent from the ISDM, Hugo Loi and Leo Giorgis from Pixminds, the LIRMM, Reference Centre for Congenital Anomalies, Clinical Genetic Unit and the Data Sciences ward of the CHU de Montpellier for their impactful discussions and contributions.

## Author Information

### Contributions

Z.L. contributed to study design, data analysis, data interpretation, figures design and writing of the manuscript. D.M. contributed to study coordination, study design and figures design. FX.D, S.G and E.P.L contributed to data collection and data management. X.C. contributed to data management and large language models predictions. P.T. contributed to the writing of the manuscript. G.M. contributed to the ethical and legal framework of the study and writing of the manuscript. K.Y. contributed to study coordination, study design, data

interpretation, figures design and writing of the manuscript. All Authors contributed to the critical review of the manuscript.

