## Appendices for "Self-Logical Consistency Assessment of Large Language Models for Patient Feedback Classification : Algorithm Development and Validation Study"

### Supplementary Note 1 : Operational definition of categories (english translation)

#### Patient Care Process

Fluidity and customization of the pathway

- Efficiency or cost-effectiveness of the organization's management
- Fluidity of the medical care pathway
- Speed of obtaining appointments
- Difficulty accessing specialist doctors
- Diversity of examinations performed
- Number of examinations performed
- Cancellation or rescheduling of an operation, delayed surgery
- Issues with operating room scheduling
- Lack of preparation and supply of implants needed for surgery
- Personalization of care, adaptation to clinical state, patient needs and preferences
- Movement within the facility by foot or stretcher, adapted to the clinical state and patient preferences
- Mode of access to the operating room, by foot or stretcher
- Consideration and involvement of family caregivers and companions in care
- Consideration of any disability, whether physical or psychological, of the patient
- Individualized accompaniment of a child
- Lack of coordination between teams and/or services; quality of communications among healthcare professionals
- Transfer due to lack of beds or space, placement in a facility distant from the family

Reception and Admission

- Location of administrative reception
- Waiting time during administrative admission procedures
- Number of staff handling administrative admissions
- Accessibility of automated administrative admission kiosks
- Presence of waiting tickets at the administrative admission location
- Contact (friendliness, impersonality, empathy, support, availability) of staff in charge of administrative admissions
- Digital pre-administration

Administrative circuit

- Administrative procedures related to the stay
- Number and repetition of documents to fill out
- Complexity of administrative pathways
- Use of digital technology for administrative procedures
- Vital card
- Shared Medical File
- Patronymic errors related to administrative procedures
- Patient identification, identity vigilance
- Theft of belongings and security of personal effects
- Ease of access to the medical record for the patient or review of the content

Speed of care and waiting time

- Speed of intra-hospital care
- Waiting time within the facility outside admissions
- Disrespect for given schedules
- Punctuality of professionals
- Opportunity to interact with healthcare professionals during waiting times
- Access to a room during waiting times
- Access to music during waiting times
- Comfort during waiting times
- Presence of recreational activities for children during waiting times
- Presence of digital access during waiting times

Access to the Operating Room

- Specific attire for surgical care
- Comfort conditions while waiting before surgery
- Visibility of the operating room or surgical field during the wait
- Asepsis and hygiene pre and per-operative

Leaving the Facility

- Access to complete information for home return
- Information on warning signs and complications after surgery
- Waiting time to obtain the surgical report
- Access to a document (discharge letter) at the end of hospitalization
- Conditions and schedules for discharge
- Transport by ambulance or taxi
- Availability of beds and places in downstream facilities
- Contact with independent practitioners, organizations, and social services for discharge
- Anticipation and arrangements for discharge

Post-Discharge Follow-Up

- Telephone reminder the day after outpatient surgery
- Ability to manage certain administrative aspects online
- Post-hospitalization follow-up by email or phone
- Need for rehospitalization

Additional Costs and Overruns

- Cost of hospitalization for the patient, price of a private room
- Reimbursement delays
- Partial reimbursements
- Visibility on reimbursements at the time of payment
- Free healthcare
- Coverage by complementary health insurance
- Fees related to private practice (display, right to overcharge)
- Deposits, guarantees

#### Professionalism and Medical and Paramedical Care

Information and explanations

- Clear and precise explanations about a disease, treatment, or prognosis, given during a consultation with a doctor
- Explanations about risks, consequences of the disease
- Explanations about care planning
- Explanations in the patient's language
- Explanations in the form of diagrams
- Explanations in the form of a booklet for reference after a consultation
- Written instructions in addition to oral information
- Detail of oral information and reports
- Consistency of information provided by different healthcare professionals
- Explanations about complications, side effects, convalescence duration, delay before resuming work, sports, or sexual activity
- Ease of access to examination results
- Information and explanations provided in vocabulary adapted to the patient's age and disabilities
- Maintaining the quality of information despite emergency situations or non-scheduled care context
- Different practitioner between anesthesia consultation and operation, without information given to the patient
- Not being informed at the moment of anesthesia
- Lack of information when the procedure performed in the operating room is about to deviate from that discussed in consultation
- Clear and detailed information about childbirth and its aftermath
- Identification and spontaneous presentation of professionals
- Quality of communications between healthcare professionals and the patient

Humanity and the availability of professionals

- The "kindness," "compassion," "closeness," "availability," "patience," "passion for work," "dedication," listening, or "empathy" displayed by the healthcare staff
- Pleasant and adapted contact by the healthcare staff
- Moral support
- A "lack of listening," "casualness," a "condescending" attitude, or a lack of goodwill displayed by the healthcare staff
- Modalities of announcing death or worsening of a condition
- Trivialization of conditions
- Patient recognition of the difficult working conditions of healthcare professionals
- Opportunity to see the responsible physician for their care
- Opportunity to see the surgeon during the stay after surgery
- Lack of time available to care teams to attend to patients, medical or caregiving absenteeism
- Absence of a referring physician in care
- Guilt-inducing attitudes of healthcare professionals
- Brutality in the procedure or lack of prior request for consent by obstetrician-gynecologists or midwives in a maternity context
- Listening ability of the care team during the care of a child
- Serious and relaxed contact of professionals in a maternity or pediatric context
- Reassurance of children and parents during hospitalization and surgery of a child

Medical and paramedical care

- Quality and speed of regulation and response to emergency calls (EMS, emergency services)
- Speed and appropriateness of initiating transport in emergency situations
- Rapid care in intra-hospital emergency situations
- Rapid decision-making between examinations
- Challenging of a professional judgment deemed wrong or failing requiring rehospitalization
- Accusation of violating medical secrecy, respect for professional secrecy, confidentiality
- Professional ethics
- Qualifications, skills of professionals
- Absence of auscultation in consultation
- Infection associated with care
- Need for re-intervention
- Confusion over the organ to be operated on
- Effectiveness of care, effectiveness of interventions
- Presence of the expression: "[…] X saved my life," designating a particular professional or an institution
- Problems with infusion or catheter placement for a peripheral venous line
- Enhanced rehabilitation after surgery
- Surgical care
- Blood pressure measurement (taking constants) at night
- Lack of monitoring
- Falls
- Discharges without the team's knowledge
- Unexpected medical or surgical complications, therapeutic hazard
- Absence/insufficiency of rehabilitation
- Absence/lack of accompaniment during meals, toileting, and everyday life activities; refusal of help from a professional

Patients' rights

- Freedom to come and go
- Civil rights (voting, etc.)
- Right to redress
- Abuse, violence, serious negligence, etc.
- End-of-life care
- Advance directives and trusted person
- Racial, religious, sexual discrimination, etc.
- Information in case of care-associated damage
- Possibilities for complaints, recourse to a mediator and mediation report, information on user representatives and the user commission, etc.
- Post-mortem care, body preservation
- Respect for dignity

Pain management and medications

- Side effects of treatments
- Information about side effects
- Consideration of the patient's drug intolerances and allergies
- Prescriptions poorly adapted to the unique situations of some patients
- Use of scales to assess pain level and reevaluation
- Consideration of pain
- Waiting in a context of pain management
- Post-hospitalization pain
- Pain management during childbirth and postpartum
- Modalities of medication delivery

Maternity and pediatrics

- Agreement between the delivering mother and healthcare staff on the personal birth plan
- Opportunity to use the "natural room"
- Support for breastfeeding
- Opportunity for skin-to-skin contact
- Facilitation of the father's involvement
- Reassurance in case of doubts about the early care of the newborn
- Advice on basic practices
- Self-dosing of the epidural by the woman
- Availability of balls for preparation
- Support during postpartum recovery
- Multiple disturbances during the maternity period
- Cesarean section
- Visits and participation of the father
- Father's training in baby care
- Presence of activities for children
- Presence of clowns
- Access to toys, books, stuffed animals
- Possibility for parents to accompany their child to the operating room
- Awarding of diplomas to young patients after surgery
- Attention to difficulties related to parent-child separation during a procedure

#### Hotel Quality

Access to the facility

- Access to nearby, free parking with available and well-maintained spaces
- Access to a drop-off zone
- Suitability of exterior equipment to standards adapted for disabilities
- Access to the facility via public transportation networks
- Road signs indicating access to the facility
- In-hospital signage and information about the location of different services, including from admissions
- Procedures, premises, and visiting hours for visitors, companions, families, including COVID visits

Premises and Rooms

- Age of premises and rooms
- Cleanliness of premises and rooms
- Maintenance of room equipment
- Access to storage in rooms
- Accessibility of the room
- Access to a refrigerator in the room
- Comfort of the mattress
- Condition of the equipment
- Quality of the bed
- Quality of the linens
- Adaptation of bed sizes for tall or large individuals
- Access to chairs for visitors
- Accommodations for companions
- Age of the shower
- Length of the shower hose
- Hygiene of the shower
- Shower leaks
- Odors from the shower
- Presence of a shower curtain
- Water drainage in the shower
- Presence of a bath mat
- Accessibility of the shower adapted to the patient's physical abilities
- Access to a towel and soap
- Access to storage furniture with functional locks
- Difficulty accessing amenities for disabled individuals
- Bathtub suitable for pregnant women
- Proximity of rooms causing difficulty sleeping in a maternity or pediatric service context
- Adaptation of premises for child patients (toilet height, etc.)
- Functional arrangement of the room
- Pleasant and spacious nature of the room
- Light disturbances

Privacy

- Access to a private room
- Respect for patient privacy and personal modesty
- Separations between patients in a shared room
- Feeling of proximity
- Feeling of personal security
- Lack of privacy in the operating room, Proximity during the wait before surgery, Mixed-gender waiting areas before surgery, Number of patients per box after surgery

Noise/Sound Level

- Acoustic isolation of the establishment, of the room
- Discretion of the caregiving teams, including at night
- Sleep deprivation due to noise
- Noise from televisions in the hallways of health facilities
- Awareness of the sound environment by the caregiving teams
- Noise caused by a neighboring room
- Air conditioning noises

Room Temperature

- Thermal insulation of the room
- Appropriateness of heating level to the room temperature
- Presence of an air conditioning system during periods of high heat
- Access to appropriate blankets
- Temperature in the operating room
- Feeling of a connection between local temperatures and the onset of nasopharyngeal diseases

Meals and Snacks

- Access to fruits and vegetables
- Taste, textures, and quantity of food
- Food adapted to the patient's feeding abilities, offering of multiple meals
- Ability to make personal choices in menu composition
- Adaptation of the menu to the patient's specific diet
- Use of plastic in hospital kitchens
- Lack of consideration for food aversions and allergies
- Meal and snack times

WiFi and TV Services

- Access to television and wifi
- State of the wifi, cellular data, and telephone network in the establishment
- Rates for television and wifi services

### Supplementary Note 2 : prompts

#### LLM Standalone prompt

*(Mentions in italic are commentaries and are not provided in the prompt)*

As the quality of care manager in a French hospital, your task is to identify specific categories and the corresponding tone mentioned in a patient textual commentary (named verbatim). The verbatims and categories are written in french. The commentary to process can be written by a patient or by another hospital agent. The commentary to process is about a recent hospitalization. It is crucial to identify a category only if it is explicitly mentioned in the commentary.

The tone of a category is defined as following :

- If the category is mentioned in a positive way, as a compliment of the quality of care, the tone is "positive"
- If the category is mentioned in a negative way, as a critic of the quality of care, the tone is "negative"
- If the category is mentioned in a neutral way, as a state of the quality of care without compliment nor critic, the tone is "neutral"
- If the category is not mentioned, the tone is "not mentioned"

The categories are distributed among three themes *(English translation)*:

- “Care Pathway”: [“The fluidity and personalization of the pathway,” “Reception and admission,” “Administrative process,” “Speed of care and waiting time,” “Access to the operating room,” “Discharge from the facility,” “Follow-up after hospital stay,” “Additional fees and out-of-pocket expenses”]
- “Team Professionalism”: [“Information and explanations,” “Humanity and availability of professionals,” “Medical and paramedical care,” “Patient rights,” “Pain management and medications,” “Maternity and pediatrics”]
- “Hotel Quality”: [“Access to the facility,” “Premises and rooms,” “Privacy,” “Calm/noise level,” “Room temperature,” “Meals and snacks,” “WiFi and TV services”]

Your output must be presented as a json file, including the theme, the category, the tones and the justification of your classification. The json must be complete with all 21 categories, even if some are not mentioned. For each couple category/tone, you must justify your classification with the following syntax :

- If you identify the presence of the tones "positive","negative" or "neutral", the justification must be an explanation of your classification, followed by the symbol | , followed by the citation of the sentence in the commentary that justifies your interpretation. Here is an example of a valid syntax : "The patient indicates the speed of his hospital treatment at the emergency ward | J'ai été très vite vu par un médecin alors même que les urgences étaient bondées"
- The justifications of the tones "positive","negative" or "neutral" that you identify as absent must remain empty.
- If all the tones "positive","negative" or "neutral" are identified as absent, you must identify the tone "not mentioned" as present. The corresponding justification must be in that case "yes".
- If any of the tones "positive","negative" or "neutral" is identified as present, you must identify the tone "not mentioned" as absent. The corresponding justification must be in that case "no".

The structure of the json file to complete is the following (english translation):

{

"**Patient Care Process**": {

"Fluidity and customization of the pathway": {

"positive": "",

"negative": "",

"neutral": "",

"not mentioned": ""

}, … *(the complete json is provided in the prompt and Supplementary Note 1, see the code for details)*

Write your output by completing the json file in totality. Be precise and restrictive in your categories identification. Identify a category only if it is explicitly mentioned in the commentary. Be sure that your json output contains the 3 themes, the 21 categories, the 4 tones and a justification for each tone. Your output must contain only the json file. Do not add any extra text.

The commentary you have to classify is the following : "*[Insert patient feedback]*“

#### Logical Consistency Prompt

*(This prompt is provided in response to the LLM answer of the LLM standalone prompt.)*

A category can be identified as present only if one of its elements is mentioned. Here is a list of each possible elements for each category in a json format :

""" *[Insert Supplementary Note 1 to json format]*"""

In this list, the first layer represents the themes, the second layer represents the categories, the third layer represents the elements. The presence of an element implies that you must identify the corresponding category as present. For each couple category/tone, you must justify your classification with the following syntax :

- If you identify the presence of the tones "positive","negative" or "neutral", the justification must exactly be the element of the given list that allows the identification of its corresponding category, followed by the symbol | , followed by the citation of the sentence in the commentary that justifies your interpretation. Here is an example of a valid syntax : " Rapidité de prise en charge en situation d’urgence intra hospitalière | J'ai été très vite vu par un médecin alors même que les urgences étaient bondées"
- The justifications of the tones "positive","negative" or "neutral" that you identify as absent must remain empty.
- If all the tones "positive","negative" or "neutral" are identified as absent, you must identify the tone "not mentioned" as present. The corresponding justification must be in that case "yes".
- If any of the tones "positive","negative" or "neutral" is identified as present, you must identify the tone "not mentioned" as absent. The corresponding justification must be in that case "no".

Next is an example of a correctly filled json output for the following fictive commentary (english translation):

'Positive points: Very warm welcome and support. Friendly attention and respect of the staff towards the patients During a postoperative call I was well reassured and advised by the intern of the department. Negative points: The location of the intervention was not very precise for a person who does not know the hospital. The services mentioned are not always understandable to external patients.' :

*(Only identified categories are given here, translated in english. The prompt contains the whole json. See the code for details. Cuts are indicated by three dots ‘...’)*

{

"**Circuit de prise en charge**": {

"Post-Discharge Follow-Up": {

"positive": "Post-hospitalization follow-up by email or phone | During a postoperative call I was well reassured and advised by the intern of the department.",

"negative": "",

"neutral": "",

"not mentioned": "no"

}, …

"**Professionalism and Medical and Paramedical Care**": {

"Information and explanations": {

"positive": "Clear and precise explanations about a disease, treatment, or prognosis, given during a consultation with a doctor | During a postoperative call I was well reassured and advised by the intern of the department.",

"negative": "",

"neutral": "",

"not mentioned": "no"

},

"Humanity and the availability of professionals": {

"positive": "The "kindness," "compassion," "closeness," "availability," "patience," "passion for work," "dedication," listening, or "empathy" displayed by the healthcare staff",

"negative": "",

"neutral": "",

"not mentioned": "no"

},...

"**Hotel Quality**": {

"Access to the facility": {

"positive": "",

"negative": "In-hospital signage and information about the location of different services, including from admissions | The location of the intervention was not very precise for a person who does not know the hospital.",

"neutral": "",

"not mentioned": "no"

},... }

Write again your output for the commentary '*[Insert patient feedback]*'. Create the json in totality according to all instructions given. In the case where you identify the presence of the tone 'positive', 'negative' ou 'neutral', it is crucial that the justification contains word for word an element of the given list defining this very category.

### Supplementary Table 1 : Benchmark sub-groups analysis

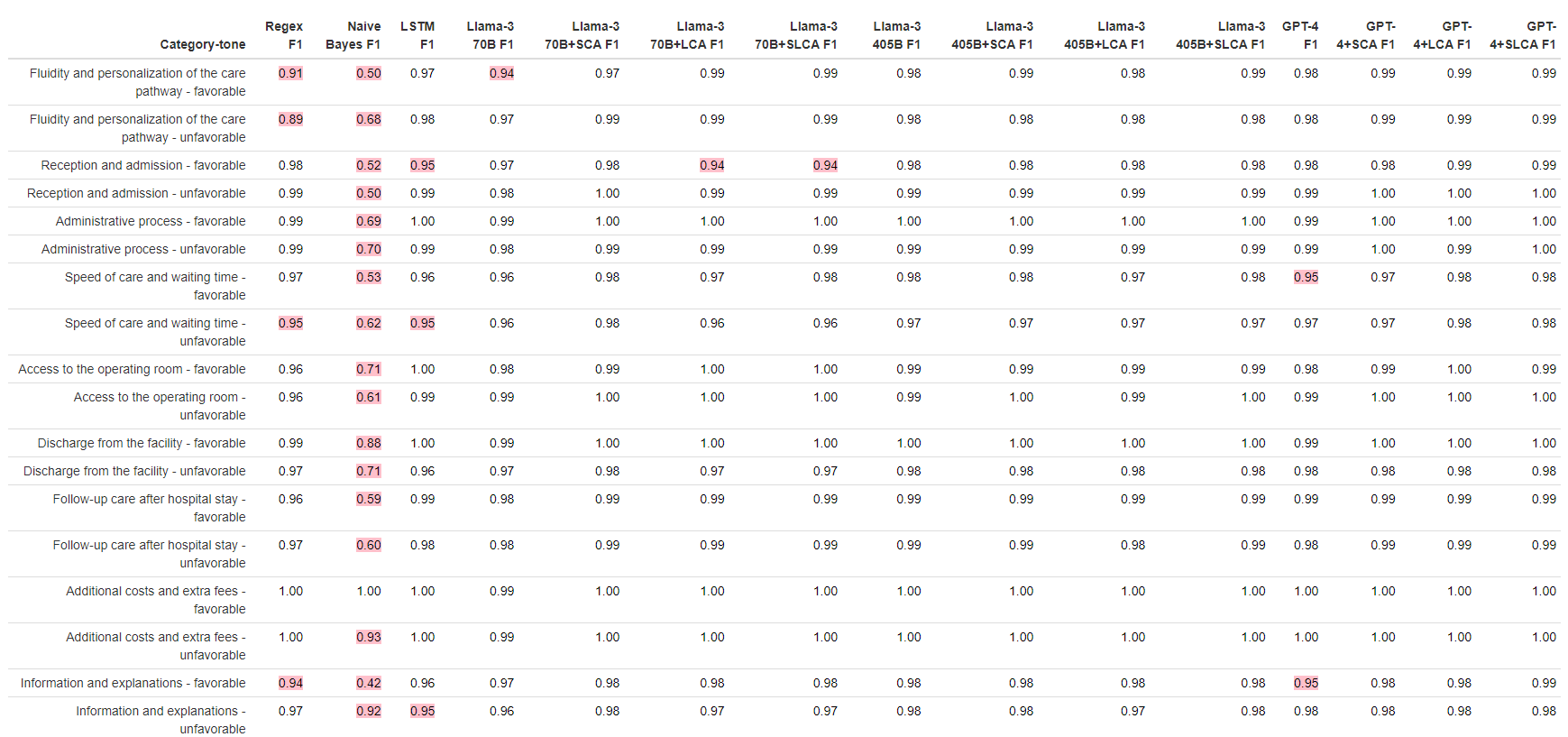

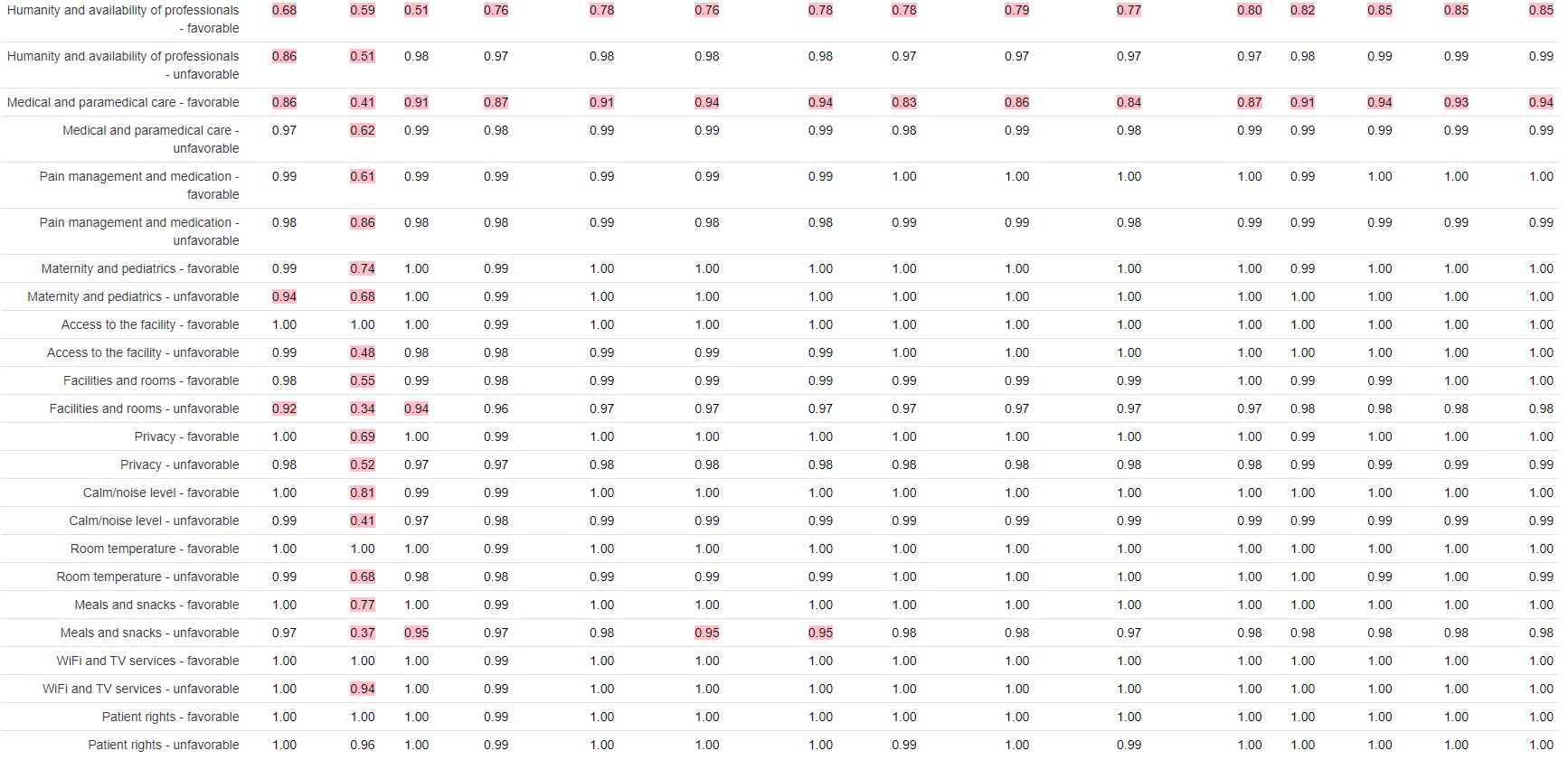

**Sub-groups performances of 9 models including LLMs+Self-Logical Consistency Assessment, classifying 1170 patient feedbacks in 42 categories**

Heuristic presented in this table are F1scores. Highlighted in red for values <=0.95.

### Supplementary Figure 1 : Flow Chart

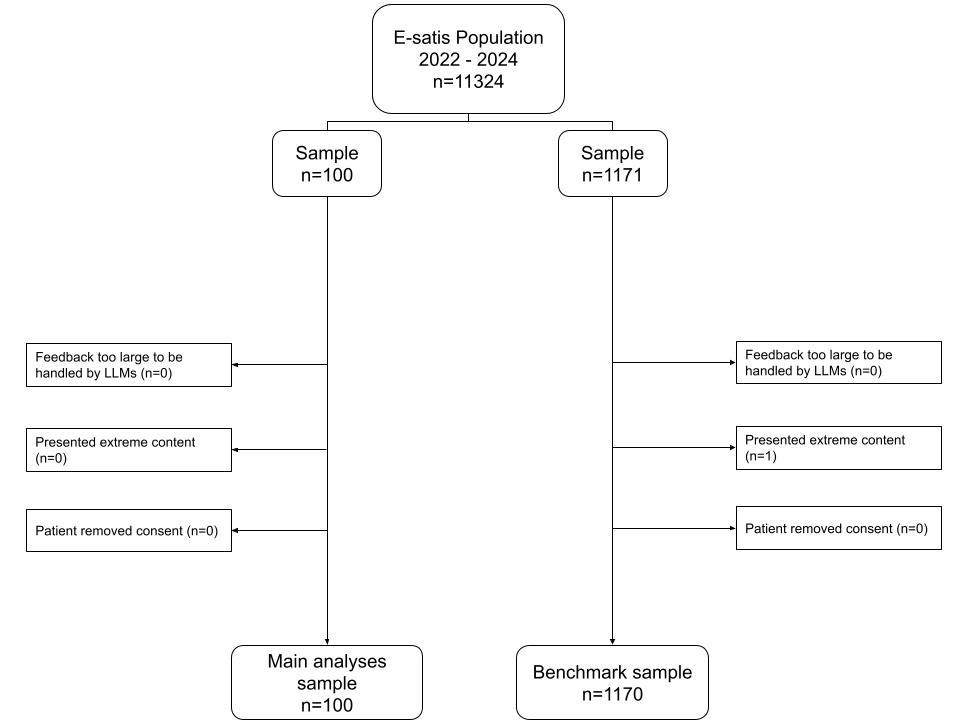

**1271 participants flow chart to study Self-Logical Consistency Assessment method in patient feedback classification**

Only one feedback has been discarded due to extreme content. A total of 1270 feedbacks from the original 1271 has been conserved.

### Supplementary Table 2 : Humans compared to GPT-4 Consistency-Assessed - Feedback sources

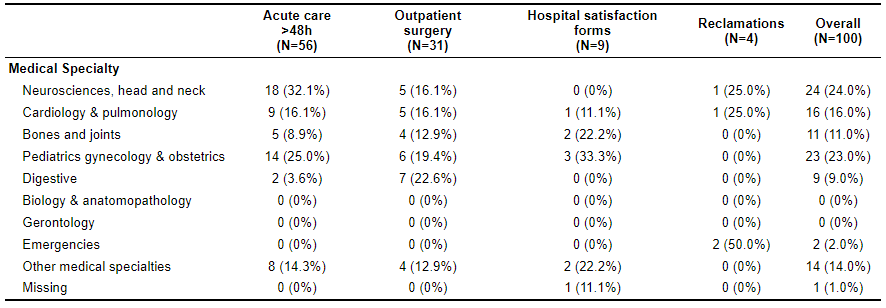

**Sources of 100 patient feedbacks to study Self-Logical Consistency Assessment method in their classification**

The following wards type are described as :

Clinical medicine : Dermatology, Pain, Hematology, Infectious diseases, Internal Medicine, Addictology, Vascular Diseases, Oncology & Palliative care

Emmbrun : Endocrinology, Metabolic Diseases, Burns, Kidney, Urology & Nephrology

MSO : Medicine, surgery and obstetrics

### Supplementary Table 3 : Benchmark - Feedback sources

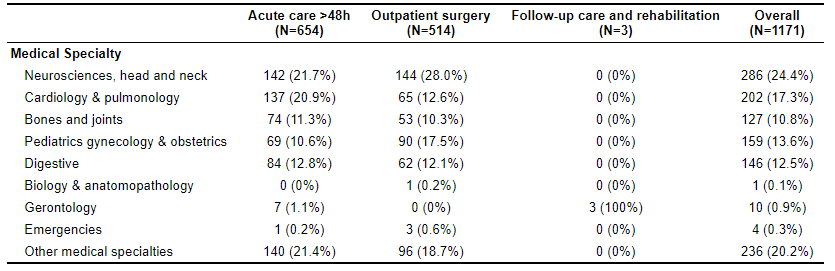

**Sources of 1170 patient feedbacks to study Self-Logical Consistency Assessment method in their classification**

The following wards type are described as :

Clinical medicine : Dermatology, Pain, Hematology, Infectious diseases, Internal Medicine, Addictology, Vascular Diseases, Oncology & Palliative care

Emmbrun : Endocrinology, Metabolic Diseases, Burns, Kidney, Urology & Nephrology

MSO : Medicine, surgery and obstetrics

### Supplementary Table 4 : Humans compared to GPT-4 Consistency-Assessed Analyses - Gold standard categories to identify

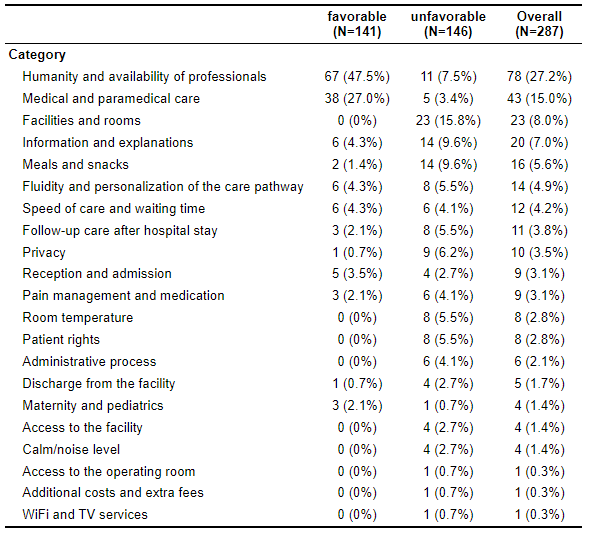

**Gold standard content of 100 patient feedbacks to study Self-Logical Consistency Assessment method in their classification**

### Supplementary Table 5 : Benchmark - Gold standard categories to identify

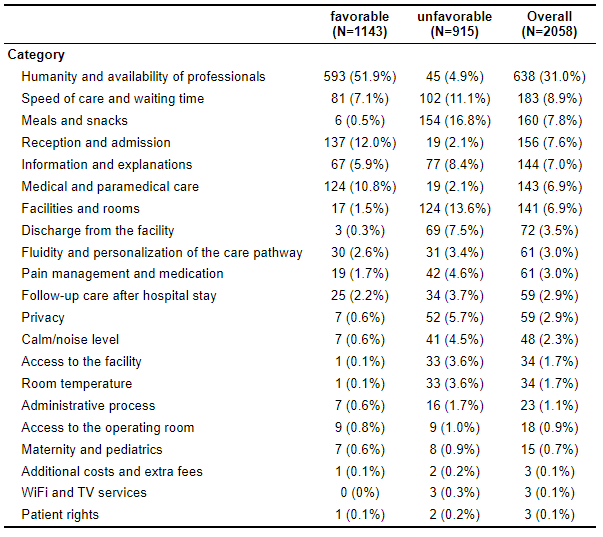

**Gold standard content of 1170 patient feedbacks to study Self-Logical Consistency Assessment method in their classification**
